# Reducing emergency department burden through the implementation of femoral nerve block in patients with femoral fractures

**DOI:** 10.1101/2025.05.30.25328613

**Authors:** Kuo-Chen Chung, Hsiu-E Chen, Chia-Hung Yeh, Subeq Yi-Maun, Shih Chieh Yang, Ching-Hui Shen, Chih-Cheng Wu

**Author notes:** Correspondence (CCW).

## Abstract

**Background:** Timely and effective pain management is essential in the emergency department, particularly for patients with femoral fractures. Despite their potential benefits, femoral nerve blocks remain underutilized compared to intravenous opioid administration.

**Methods:** This retrospective study evaluated patients with femoral fractures who received either femoral nerve blocks (Group 1) or standard Intravenous opioid treatment (Group 2). Pain reduction was assessed using the Numerical Rating Scale, and Intravenous opioid use was recorded.

**Results:** Group 1 experienced a significantly greater reduction in pain scores (mean decrease of 4.96 vs. 3.99; p < 0.001). While the frequency of Intravenous opioid use was slightly lower in Group 1 (1.35 vs. 1.64; p = 0.063), the trend suggests a reduction in opioid reliance. The average time to additional opioid administration post-femoral nerve blocks was 6.49 hours.

**Conclusions:** Femoral nerve blocks offer superior analgesia for femoral fractures and may reduce opioid use in the emergency departments, potentially easing the burden on emergency staff. Further studies are warranted to validate these findings.

## 1. Introduction

### 1.1 Background and Problem Statement

Femoral fractures, particularly those involving the hip, are among the most painful injuries encountered in emergency departments (ER), disproportionately affecting the elderly. Effective pain management in these patients is crucial not only for comfort but also for preventing complications such as delirium, reduced mobility, and extended hospital stays. Intravenous opioid (IVO) analgesia is traditionally used for acute pain control in these settings; however, its efficacy is often compromised by ER overcrowding, which can lead to delays in administration and under-treatment. Moreover, opioids carry notable risks, including respiratory depression and cognitive impairment, especially in older adults.

### 1.2 Emerging Alternatives

Femoral nerve blocks (FNB) have emerged as a promising alternative, offering targeted analgesia with fewer systemic side effects. Despite their advantages, FNB remain underutilized in emergency care due to variability in training, procedural complexity, and institutional limitations.

### 1.3 Literature Overview

Recent studies support the clinical effectiveness of FNBs. For example, Scurrah et al. demonstrated enhanced early analgesia in elderly hip fracture patients [1], while Tsai et al. confirmed the feasibility of ultrasound-guided FNB in ED with better pain control than systemic opioids [2]. A meta-analysis by Hayashi et al. involving over 4,700 participants found that peripheral nerve blocks, including FNB, significantly reduced pain scores within two hours [3].

In addition to pain control, FNB may reduce opioid consumption and improve outcomes. Beaudoin et al. reported decreased need for rescue analgesia and fewer side effects with FNB [4]. Similarly, Gerlier et al. found early FNB reduced preoperative opioid requirements [5]. Unneby et al. further supported its use in elderly and cognitively impaired populations [6].

### 1.4 Research Gap

Despite robust evidence supporting FNB efficacy, their impact on ER workflow and opioid usage patterns is less well understood. Ritcey et al. identified barriers to FNB adoption, including lack of standardization and operational concerns[7]. Furthermore, few studies have examined whether FNBs reduce the frequency of opioid dosing and associated monitoring, potentially easing the burden on emergency staff.

### 1.5 Study Aim

To address this gap, the current study retrospectively analyzes patients with femoral fractures who received either FNB (Group 1) or standard IVO treatment (Group 2). Outcomes assessed include changes in pain scores using the Numerical Rating Scale (NRS), frequency of opioid administration, time to additional analgesia, and implications for ER workload. The findings aim to inform clinical practice by evaluating both the therapeutic and operational benefits of FNB implementation in emergency care.

## 2. Materials and Methods

### 2.1. Study Design and Setting

This retrospective comparative study was conducted at the ER of Taichung Veterans General Hospital between April 2021 and September 2023. The primary aim was to assess the analgesic effectiveness and opioid-sparing potential of FNB compared to standard IVO treatment in patients with femoral fractures.

Pain severity was measured using an 11-point NRS, ranging from 0 (no pain) to 10 (worst imaginable pain). NRS scores were recorded immediately before and 60 minutes after the pain control intervention (PCI). All NRS assessments were performed by trained nursing staff who were independent of the physicians performing the interventions, minimizing potential observer bias.

Eligible participants were adults (aged ≥ 18 years) who presented to the ER with radiologically confirmed femoral fractures—including fractures of the femoral head, neck, intertrochanteric region, shaft, or distal femur—and an initial NRS score of ≥ 7. Patients were excluded if they had cognitive impairment, multiple or pathological fractures, known coagulopathies, infection at the intended injection site, hypersensitivity to local anesthetics, unstable vital signs, altered mental status, pregnancy or lactation, or a body mass index > 30 kg/m2.

#### Patients were assigned to one of two groups

Group 1 (FNB group): Patients who received a femoral nerve block administered by a single experienced emergency physician using ultrasound guidance.

Group 2 (IVO group): Patients who received standard pain management consisting of intravenous tramadol at 1 mg/kg.

To reduce selection bias, an equal number of patients were included in each group, drawn from the same study period. Upon diagnosis, the hospital information system notified the physician for immediate intervention. If NRS scores remained ≥ 7 after PCI, a rescue IVO dose was administered.

The following parameters were collected: patient demographics, fracture site, Injury Severity Score (ISS), triage level, number of IVO administrations during the ER stay, emergency room stay time (ERS, in hours), the NRS before and after PCI, and the magnitude of NRS reduction.

### 2.2. FNB Procedure Protocol

All FNB were performed using a high-frequency linear ultrasound probe (8–18 MHz) to identify anatomical landmarks including the common femoral artery, iliopsoas muscle, sartorius muscle, and fascia iliaca. Using an in-plane approach, a 21-gauge needle was advanced under visualization to deposit local anesthetic (20 mL of 0.25% bupivacaine) adjacent to the femoral nerve, typically located just below the inguinal ligament. Aspiration was performed after every 5 mL injection to prevent intravascular administration.

### 2.3. Data Analysis

All data were anonymized and analyzed using SPSS version 27.0 (IBM, Armonk, NY). Categorical variables were reported as frequencies and percentages, while continuous variables were expressed as means with standard deviations (SD) or medians where appropriate. Normality of distribution was assessed using the Shapiro–Wilk test.

Group comparisons for categorical variables were performed using Chi-square or Fisher’s exact tests. Continuous variables were analyzed using the independent samples t-test or the Mann–Whitney U test, depending on data distribution. Pearson’s correlation coefficients were calculated to assess associations among continuous variables. Both univariate and multivariable linear regression analyses were conducted to explore predictors of opioid use and pain score reduction. A p-value of <0.05 was considered statistically significant.

### 2.4. Outcome Measures

The primary outcome was the NRS reduction 60 minutes after the intervention, comparing FNB with IVO treatment. The secondary outcome was the number of opioid administrations required during the ER stay. Adverse events, including nerve block-related complications and any systemic reactions, were also documented.

## 3. Results

### 3.1. Baseline Characteristics

A total of 248 patients were included in the analysis: 134 in the FNB group (Group 1) and 114 in the IVO group (Group 2). As shown in Table 1, there were no statistically significant differences between the groups in terms of age (68.90 ± 23.11 vs. 66.06 ± 22.43 years, p = 0.330), sex distribution (female: 59.0% vs. 64.0%, p = 0.413), triage level (p = 0.423), or femoral fracture location (p = 0.195). These results indicate that the two groups were comparable at baseline.

**Table 1.** N=248, Demographics and Baseline Characteristics of Study Participants.

|  | Group1 (n=134) | Group2 (n=114) | *p* value |
| --- | --- | --- | --- |
| Sex |  |  | 0.413 |
| Female | 79 (59.0%) | 73 (64.0%) |  |
| Male | 55 (41.0%) | 41 (36.0%) |  |
| Age | 68.90 ±23.11 | 66.06 ±22.43 | 0.330 |
| triage |  |  | 0.423 |
| 1 | 2 (1.5%) | 3 (2.6%) |  |
| 2 | 33 (24.6%) | 21 (18.4%) |  |
| 3 | 99 (73.9%) | 90 (79.0%) |  |
| femoral fracture location |  |  | 0.195 |
| upper third | 87 (64.9%) | 84 (73.7%) |  |
| middle third | 29 (21.6%) | 22 (19.3%) |  |
| low third | 18 (13.4%) | 8 (7.0%) |  |

Chi-square test or Independent t test, Mean (SD). \* $p < 0.05$

### 3.2. Pain Reduction and Opioid Use

Table 2 summarizes pain scores and opioid use during ERS. The NRS before PCI were similar between groups (7.96 ± 0.59 in Group 1 vs. 8.04 ± 0.69 in Group 2; p = 0.375). However, Group 1 exhibited a significantly greater reduction in pain following intervention. The mean NRS after PCI was 3.00 ± 1.21 in Group 1 compared to 4.04 ± 2.28 in Group 2 (p < 0.001), with corresponding NRS reductions of 4.96 ± 1.22 and 3.99 ± 2.26, respectively (p < 0.001).

**Table 2.**

|  | Group1 (n=134) | Group2 (n=114) | p value |
| --- | --- | --- | --- |
| ERS (hour) | $13.56 \pm 8.54$ | $11.98 \pm 7.11$ | 0.114 |
| How many times IVO during ERS | $1.35 \pm 1.37$ | $1.64 \pm 1.01$ | 0.063 |
| NRS before PCI | $7.96 \pm 0.59$ | $8.04 \pm 0.69$ | 0.375 |
| NRS after PCI | $3.00 \pm 1.21$ | $4.04 \pm 2.28$ | $<0.001^{**}$ |
| NRS reduction | $4.96 \pm 1.22$ | $3.99 \pm 2.26$ | $<0.001^{**}$ |

Chi-square test or Independent t test, Mean (SD). \* $p < 0.05$
ERS: emergency room stay time; IVO: intravenous opioid; NRS: numerical rating scale; PCI: pain control intervention

While the average number of IVO administrations during ERS was lower in Group 1 (1.35 ± 1.37) than in Group 2 (1.64 ± 1.01), the difference approached but did not reach statistical significance (p = 0.063). Importantly, only 49 patients (36.6%) in Group 1 required additional IVO after FNB during ERS. The average time to the first rescue opioid dose in this subgroup was 6.49 hours, with the maximum delay extending up to 18.8 hours. ERS were comparable between groups (13.56 ± 8.54 hours for Group 1 vs. 11.98 ± 7.11 hours for Group 2; p = 0.114).

### 3.3. Correlation Analysis

Table 3 presents Pearson correlation coefficients for key variables across all patients (n = 248). Several notable relationships emerged:

Age and Triage Severity: Older patients tended to present with more severe triage levels (r = 0.347, p < 0.01).

Pain Control and Opioid Frequency: A negative correlation was found between NRS reduction and the frequency of IVO administration (r = –0.201, p < 0.01), suggesting that inadequate initial pain control may lead to repeated opioid dosing.

Pain Scores and Pain Improvement: A strong inverse correlation between post-intervention NRS and NRS reduction (r=–0.940, p < 0.01) underscores the effectiveness of pain management in the FNB group.

Opioid Use and ERS: ERS time was positively correlated with opioid administration frequency (r=0.460, p<0.01), indicating that patients who required more analgesics tended to remain in the ER longer.

**Table 3.**
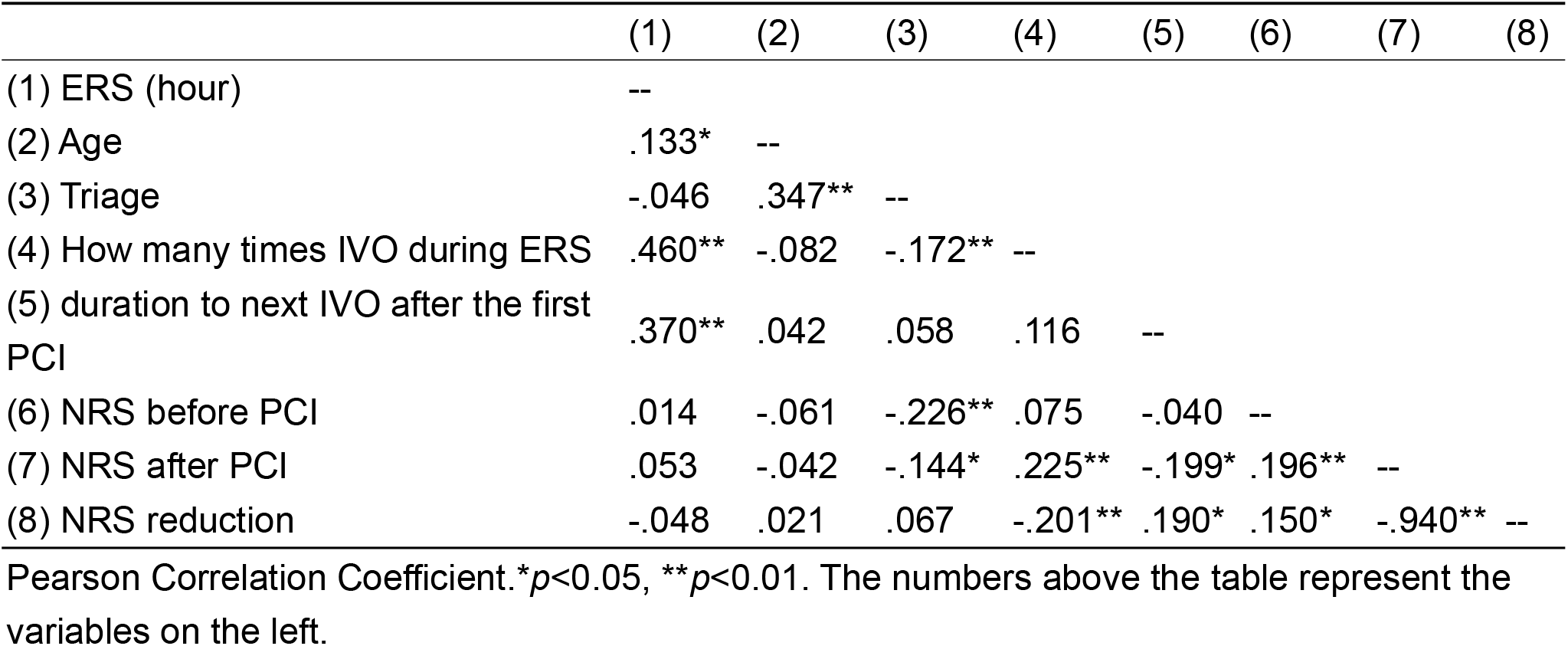

|  | (1) | (2) | (3) | (4) | (5) | (6) | (7) | (8) |
| --- | --- | --- | --- | --- | --- | --- | --- | --- |
| (1) ERS (hour) | -- |  |  |  |  |  |  |  |
| (2) Age | .133* | -- |  |  |  |  |  |  |
| (3) Triage | -.046 | .347** | -- |  |  |  |  |  |
| (4) How many times IVO during ERS | .460** | -.082 | -.172** | -- |  |  |  |  |
| (5) duration to next IVO after the first PCI | .370** | .042 | .058 | .116 | -- |  |  |  |
| (6) NRS before PCI | .014 | -.061 | -.226** | .075 | -.040 | -- |  |  |
| (7) NRS after PCI | .053 | -.042 | -.144* | .225** | -.199* | .196** | -- |  |
| (8) NRS reduction | -.048 | .021 | .067 | -.201** | .190* | .150* | -.940** | -- |

Pearson Correlation Coefficient. \* $p < 0.05$ , \*\* $p < 0.01$ . The numbers above the table represent the variables on the left.

## 4. Discussion

Femoral fractures, particularly those involving the hip, represent one of the most significant orthopedic emergencies, especially in elderly populations. These injuries are frequently associated with high levels of acute pain, functional decline, and increased morbidity. In older adults, unmanaged or inadequately managed pain can exacerbate frailty and is linked to a cascade of adverse events, including delirium, reduced mobility,hospital-acquired complications, longer hospital stays, and elevated mortality rates [8-10]. Effective pain control in this context is not merely a comfort measure but a cornerstone of early intervention to improve both immediate and long-term outcomes.

Traditionally, IVO have been the standard approach for managing fracture-related pain in the ED. However, opioids come with well-documented risks, such as respiratory depression, nausea, vomiting, hypotension, and especially delirium in geriatric patients [7, 11, 12]. Moreover, their administration in a busy ER setting often involves complex workflows, frequent monitoring, and the potential for medication delays or underdosing. These challenges underline the need for alternative analgesic strategies that are both clinically effective and operationally efficient. Some studies have shown that the use of peripheral nerve blocks and multimodal analgesia techniques may reduce the incidence of delirium and facilitate early mobilization, thus ultimately reducing morbidity and post-op 1-year mortality after hip surgery[11, 13].

In this study, we investigated whether the early application of ultrasound-guided FNB could serve as a viable alternative to IVO administration for patients presenting to the ER with femoral fractures. Specifically, we sought to determine whether FNBs not only provide superior pain relief but also reduce the reliance on opioids and the burden on emergency department resources.

### 4.1. Primary Outcome: Pain Reduction

Our findings indicate a clear and statistically significant advantage of FNB over standard IVO therapy in terms of acute pain reduction. The mean reduction in the NRS score was substantially greater in the FNB group (4.96) compared to the IVO group (3.99), with a p-value of < 0.001, denoting strong statistical significance. Furthermore, the post-intervention NRS score in the FNB group dropped to a mean of 3.00, which is within the range of mild pain, whereas the IVO group remained at a higher average of 4.04 (p < 0.001). This difference is not only statistically relevant but also clinically meaningful, as crossing the threshold from moderate to mild pain has implications for both patient comfort and downstream care processes.

The superior efficacy of FNB likely stems from its localized mechanism of action. By delivering anesthetic directly to the femoral nerve, FNB provides targeted, rapid, and sustained analgesia that is well-suited to the pathophysiology of femoral fractures. In contrast, IVOs work systemically and may suffer from delayed onset and broader side effect profiles. Moreover, FNB’s ability to deliver consistent pain relief without sedative effects makes it particularly attractive in older patients at higher risk of opioid-induced complications.

### 4.2. Secondary Outcome: Opioid Use and Operational Impact

In addition to superior analgesia, the use of FNB was associated with a reduction in opioid exposure during the ER stay. Only 36.6% of patients in the FNB group required rescue IVO, and the average time to additional opioid administration was significantly delayed (6.49 hours), with some patients remaining opioid-free for over 18 hours. This delay and reduced frequency of administration are particularly relevant in the current healthcare context, where concerns over opioid misuse, dependency, and side effects have become critical public health issues.

Although the difference in IVO administration frequency between groups (1.35 vs. 1.64 times) did not reach conventional statistical significance (p = 0.063), the trend suggests a clinically meaningful reduction in the need for repeat dosing. In a high-throughput ER environment, even modest reductions in medication handling, reassessment, and monitoring can translate into considerable operational benefits. Notably, correlation analysis supported this interpretation, showing that greater pain relief, as measured by NRS reduction, was associated with fewer IVO administrations. Likewise, longer ER stays were positively correlated with higher opioid usage, highlighting the compounding effects of inadequate pain control on patient flow and resource allocation.

From a systems-level perspective, these findings suggest that integrating FNB into routine ER protocols could lead to streamlined workflows, reduced medication burden, and more efficient patient throughput—all without compromising safety or efficacy.

### 4.3. Comparison with Existing Literature

The findings from our study are broadly consistent with and extend those of earlier investigations into the benefits of peripheral nerve blocks for hip and femoral fractures. Hayashi et al., in a large meta-analysis of 63 randomized trials, demonstrated that peripheral nerve blocks—including femoral, fascia iliaca, and pericapsular nerve group (PENG) blocks—provided significantly better pain relief two hours after administration compared to standard care, although effects on morphine consumption were more variable[3]. Geizhals et al. reported a reduction in preoperative morphine equivalents in patients receiving FNB, albeit without reaching statistical significance (10.3 vs. 14.0 MMEs, P = 0.13)[14]. Similarly, Häusler et al. found that regional anesthesia techniques were associated with decreased postoperative opioid requirements and improved safety profiles[15].

Importantly, studies such as those by Beaudoin and Gerlier have shown that FNB not only improves pain control but also reduces the incidence of opioid-related adverse events and the need for rescue analgesia—mirroring our own findings[4, 5]. Of particular note, Unneby et al. found that even elderly patients with cognitive impairment, such as dementia, benefited from FNBs with lower pain scores and decreased opioid consumption, emphasizing the potential of this technique in vulnerable populations [6].

### 4.4. Clinical Implications

FNB offers a practical and efficient alternative to systemic opioids in managing femoral fracture pain. Its implementation may reduce medication-related complications, minimize the frequency of pain reassessments, and alleviate the workload on ER staff—a significant benefit in high-pressure, resource-constrained environments. Identifying patient profiles that respond optimally to FNB could further enhance personalized pain management protocols and reduce unnecessary opioid exposure.

### 4.5. Study Limitations

This study has several limitations. First, its retrospective design introduces the potential for selection bias and limits causal inference. Second, the procedures were conducted at a single center by a single experienced physician, which may restrict generalizability to other settings or practitioners with varying skill levels. Third, while we measured immediate pain relief and opioid usage, we did not assess longer-term outcomes such as functional recovery, complication rates, or patient satisfaction. Lastly, potential confounders—such as pre-existing pain tolerance, prior opioid exposure, or fracture severity—were not adjusted for in the current analysis.

### 4.6. Future Directions

Further prospective, multicenter studies with randomized designs are warranted to validate these findings. Future research should also explore long-term outcomes, predictors of FNB effectiveness, and cost-effectiveness analyses to support policy-level decisions for integrating FNB into routine emergency care.

## Institutional Review Board Statement

The study was conducted in accordance with the Declaration of Helsinki, and approved by the Institutional Review Board of Taichung Veterans General Hospital (No. CE22273B-1, 18 July 2024).

## Informed Consent Statement

Patient consent was waived because this study is a retrospective comparative analysis and does not involve any identifiable participating patients.

## Data Availability Statement

Data sharing is not applicable.

## Acknowledgments

The authors thank the Biostatistics Group, Department of Medical Research, Taichung Veterans General Hospital and Mr. Chen, Jun-Peng for statistical analysis.

## Abbreviations

The following abbreviations are used in this manuscript:

NRS: numerical rating scale
IVO: intravenous opioid
ER: emergency department
ERS: emergency room stay time
FNB: femoral nerve blocks
PCI: pain control intervention
ISS: Injury Severity Score

## References

1. Scurrah A, Shiner CT, Stevens JA, Faux SG. Regional nerve blockade for early analgesic management of elderly patients with hip fracture - a narrative review. Anaesthesia. 2018;73(6):769–83.

2. Tsai TY, Cheong KM, Su YC, Shih MC, Chau SW, Chen MW, et al. Ultrasound-Guided Femoral Nerve Block in Geriatric Patients with Hip Fracture in the Emergency Department. J Clin Med. 2022;11(10).

3. Hayashi M, Yamamoto N, Kuroda N, Kano K, Miura T, Kamimura Y, et al. Peripheral Nerve Blocks in the Preoperative Management of Hip Fractures: A Systematic Review and Network Meta-Analysis. Ann Emerg Med. 2024;83(6):522–38.

4. Beaudoin FL, Haran JP, Liebmann O. A comparison of ultrasound-guided three-in-one femoral nerve block versus parenteral opioids alone for analgesia in emergency department patients with hip fractures: a randomized controlled trial. Acad Emerg Med. 2013;20(6):584–91.

5. Gerlier C, Mijahed R, Fels A, Bekka S, Courseau R, Singh AL, et al. Effect of early ultrasound-guided femoral nerve block on preoperative opioid consumption in emergency patients with hip fracture: a randomized trial. Eur J Emerg Med. 2024;31(1):18–28.

6. Unneby A, Svensson O, Gustafson Y, Olofsson B. Femoral nerve block in a representative sample of elderly people with hip fracture: A randomised controlled trial. Injury. 2017;48(7):1542–9.

7. Ritcey B, Pageau P, Woo MY, Perry JJ. Regional Nerve Blocks For Hip and Femoral Neck Fractures in the Emergency Department: A Systematic Review. Cjem. 2016;18(1):37–47.

8. Dhanwal DK, Siwach R, Dixit V, Mithal A, Jameson K, Cooper C. Incidence of hip fracture in Rohtak district, North India. Arch Osteoporos. 2013;8(0):135.

9. Marks R. Hip fracture epidemiological trends, outcomes, and risk factors, 1970-2009. Int J Gen Med. 2010;3:1–17.

10. Parikh K, Kandemir U, Agarwal A. Intertrochanteric Hip Fractures: Pearls and Pitfalls in Managing Difficult Fractures. Instr Course Lect. 2023;72:375–87.

11. Downey C, Kelly M, Quinlan JF. Changing trends in the mortality rate at 1-year post hip fracture - a systematic review. World J Orthop. 2019;10(3):166–75.

12. Lee LA, Caplan RA, Stephens LS, Posner KL, Terman GW, Voepel-Lewis T, et al. Postoperative opioid-induced respiratory depression: a closed claims analysis. Anesthesiology. 2015;122(3):659–65.

13. Ackermann L, Schwenk ES, Lev Y, Weitz H. Update on medical management of acute hip fracture. Cleve Clin J Med. 2021;88(4):237–47.

14. Geizhals S, Shou Y, Rudnin S, Tama M, Greenstein J, Hahn B, et al. Femoral nerve blocks versus standard pain control for hip fractures: a retrospective comparative analysis. Clin Exp Emerg Med. 2024;11(2):181–7.

15. Häusler G, van der Vet PCR, Beeres FJP, Kaufman T, Kusen JQ, Poblete B. The impact of loco-regional anaesthesia on postoperative opioid use in elderly hip fracture patients: an observational study. Eur J Trauma Emerg Surg. 2022;48(4):2943–52.

